# The prevalence and influencing factors for anxiety in medical workers fighting COVID-19 in China: A cross-sectional survey

**DOI:** 10.1101/2020.03.05.20032003

**Authors:** Chen-Yun Liu, Yun-zhi Yang, Xiao-Ming Zhang, Xinying Xu, Qing-Li Dou, Wen-Wu Zhang

## Abstract

**Background:** The COVID-19 outbreak caused by the SARS-Cov-2 virus has been sustained in China since December 2019, and could become a pandemic if we do not contain it. The mental health of frontline medical staff is a concern. In this study, we aimed to identify the influencing factors on medical worker anxiety in China during the COVID-19 outbreak.

**Methods:** We conducted a cross-sectional study to estimate the prevalence of anxiety among medical staff from 10^th^ February 2020 to 20^th^ February 2020 in China using the Zung Self-rating Anxiety Scale (SAS) to assess anxiety, using the criteria of normal (≤49), mild (50-59), moderate (60- 70) and severe anxiety (≥70). We used multivariable linear regression to determine the factors (e.g., having direct contact treating infected patients, being a medical staff worker from Hubei province, being a suspect case) for anxiety. We also used adjusted models to confirm independent factors for anxiety after adjusting for gender, age, education and marital status.

**Results:** Of 512 medical staff from China, 164 healthcare workers (32.03%) had had direct contact by treating infected patients. The prevalence of anxiety was 12.5%, with 53 workers suffering from mild (10.35%), seven workers from moderate (1.36%) and four workers from severe anxiety (0.78%). After adjusting for sociodemographic characteristics (gender, age, education and marital status), medical staff who had had direct contact treating infected patients saw higher anxiety scores than those who had not had direct contact (βvalue=2.33, CI: 0.65 −4.00; p=0.0068). Similar things were observed in medical staff from Hubei province, compared with those from other parts of China (β value=3.67, CI: 1.44 −5.89; p=0.0013). The most important variable was suspect cases with high anxiety scores, compared to non-suspect cases (βvalue=4.44, CI: 1.55 −7.33; p=0.0028).

**Conclusion:** Our results highlight that government authorities should make early detection of the high risk of anxiety among medical staff a priority, and implement appropriate psychological intervention programs, to prevent medical staff from developing psychological disorders that could potentially exert an adverse effect on combating the COVID-19 epidemic.

## Introduction

The COVID-19 outbreak caused by SARS-Cov-2 emerged in Wuhan, China, spread to the entire country from the end of December 2019, and has attracted enormous concern from around the world^1^. It is estimated that the number of infected patients is more than 77,043, with 2,445 deaths, which is more serious than SARS, a similar epidemic disease. The Chinese government has implemented numerous measures, including quarantines, reducing the use of public transportation, and temporarily canceling work and school, to control this disease.^2^ In addition, government authorities have sent the majority of medical staff from each Chinese province to fight COVID-19.

COVID-19 is characterized by complexity, including human-to-human transmission, asymptomatic carrier transmission and high transmission efficiency, which could lead to a worldwide pandemic^3,4^. Medical staff are frontline workers to treat infected patients, however, with a higher risk of exposure. Current source data have presented the proportion of infected medical staff at 3.8%, mainly due to early non-protected contact with infected patients^5,6^. Several previous studies reported that medical staff might suffer adverse psychological disorders, such as anxiety, fear and stigmatization, which occurred during the SARS and Ebola outbreaks, and could exert an adverse effect on care quality^7-10^. Medical staff must wear heavy protective garments and an N95 mask, making it much more difficult to carry out medical operations or procedures than under normal conditions. These factors, together with the fear of being contagious and infecting others, could increase the possibility of psychological issues among medical staff. David Koh and colleagues found that more than half of clinical staff reported increased work stress (56%) and workload (53%) during the SARS epidemic in Singapore^11^. In addition, a Hong Kong study found that health workers suffered high anxiety scores after directly treating confirmed SARS patients^12^. Therefore, it is very important to study medical workers’ mental health status. Studies exploring the prevalence of anxiety among medical staff during the COVID-19 outbreak in China are limited. Our study’s aim was to examine the anxiety levels of frontline health care workers and identify the risk factors for anxiety in China during the COVID-19 epidemic. Our findings might help governments or health authorities to recognize the causes of increased anxiety in healthcare workers, and then to provide early effective measures to reduce that anxiety.

## Methods

This is a descriptive quantitative cross-sectional study that was used to explore the prevalence and factors linked to anxiety in frontline medical staff. Data were collected from 10^th^ February 2020 to 20^th^ February 2020 in China during the COVID-19 epidemic. Informed consent was provided by subjects before study commencement. After that, we distributed self-reported questionnaires to healthcare workers via WeChat.

### Study participants

Participating healthcare staff included doctors, nurses and administrative workers at hospitals in China.

### Materials

The questionnaire consisted of three sections: (1) Demographic characteristics, such as gender, age, marital status, level of education, department, city and location (2) Questions included the following: (e.g. Have you ever directly treated a patient with COVID-19?; Have you been to Hubei province in the last month?; Are you a suspect case who had direct contact with a confirmed case or do you have a fever, fatigue, cough?; Did you adhere to the preventive and control measures in your community?; Do you need psychological treatment?) (3) The Zung Self-rating Anxiety Scale (SAS)^13^ was used to assess medical staff anxiety levels. In SAS, there are 20 items ranked on a 1-4 scale, with total raw scores ranging from 20-80. In addition, published studies conducted in China have reported satisfactory reliability and validity^14^. The higher the score, the higher the degree of anxiety. The SAS scores were classified into four categories, including normal (≤49), mild anxiety (50-59), moderate anxiety (60-70), and severe anxiety (≥70), after standardizing the score based on raw data multiplied by 1.25. Previous studies have shown that SAS internal consistency reliability was 0.66–0.80 and the Cronbach’s alpha was 0.87^15^.

### Ethical considerations

Ethical approval from the ethics committee of the People’s Hospital of Baoan District, Shenzhen (Certificate: BYL20200202) was obtained, with written consent provided by all participants. Medical staff whose SAS scores were more than 60 points were informed of their score and a researcher contacted them to ask whether they wanted psychological treatment. If they accepted, our team helped them make an online appointment for a psychological consultation.

### Data statistical analyses

Continuous and categorical variates were summarized as mean values±standard deviation (SD) and frequency (percentage), respectively. We used the Chi-square test to identify the differences in categorical variables between groups, and the students’ t-test was used to determine the differences in continuous variables between groups. If the data showed a skewed distribution, the Kruskal Wallis H Test was used. In addition, a univariate analysis model was used to identify the relationship between risk factors and anxiety score. Finally, our paper also lists the unadjusted and adjusted multivariate linear regression analysis model. Statistically significant differences were identified as a two-sided P value < 0.05. All analyses were conducted using EmpowerStats (http://www.empowerstats.com, X&Y Solutions, Inc., Boston, MA) and software package R (http://www.r-project.org).

## Results

### Patient baseline characteristics are presented in Table 1

A total of 600 medical staff was surveyed, with 512 subjects completing the questionnaire and a dropout rate of 14.6%. The majority of the sample was female (84.57%) and the largest age group was 18-39 years old. Most sample participants were married (62.50%) and the majority of subjects had a tertiary level of education. Most health workers came from a clinical department (72.07%), with 13.28% coming to set up clinics to screen patients for fever, and 14.65% for administration. A total of 164 health workers had directly treated confirmed patients as shown in Table 1, with 14.26% of respondents coming from Hubei province, the most severely affected area. A total of 53 respondents suffered from mild anxiety (10.35%), seven from moderate anxiety (1.36%) and four from severe anxiety (0.78%).

**Table 1.**
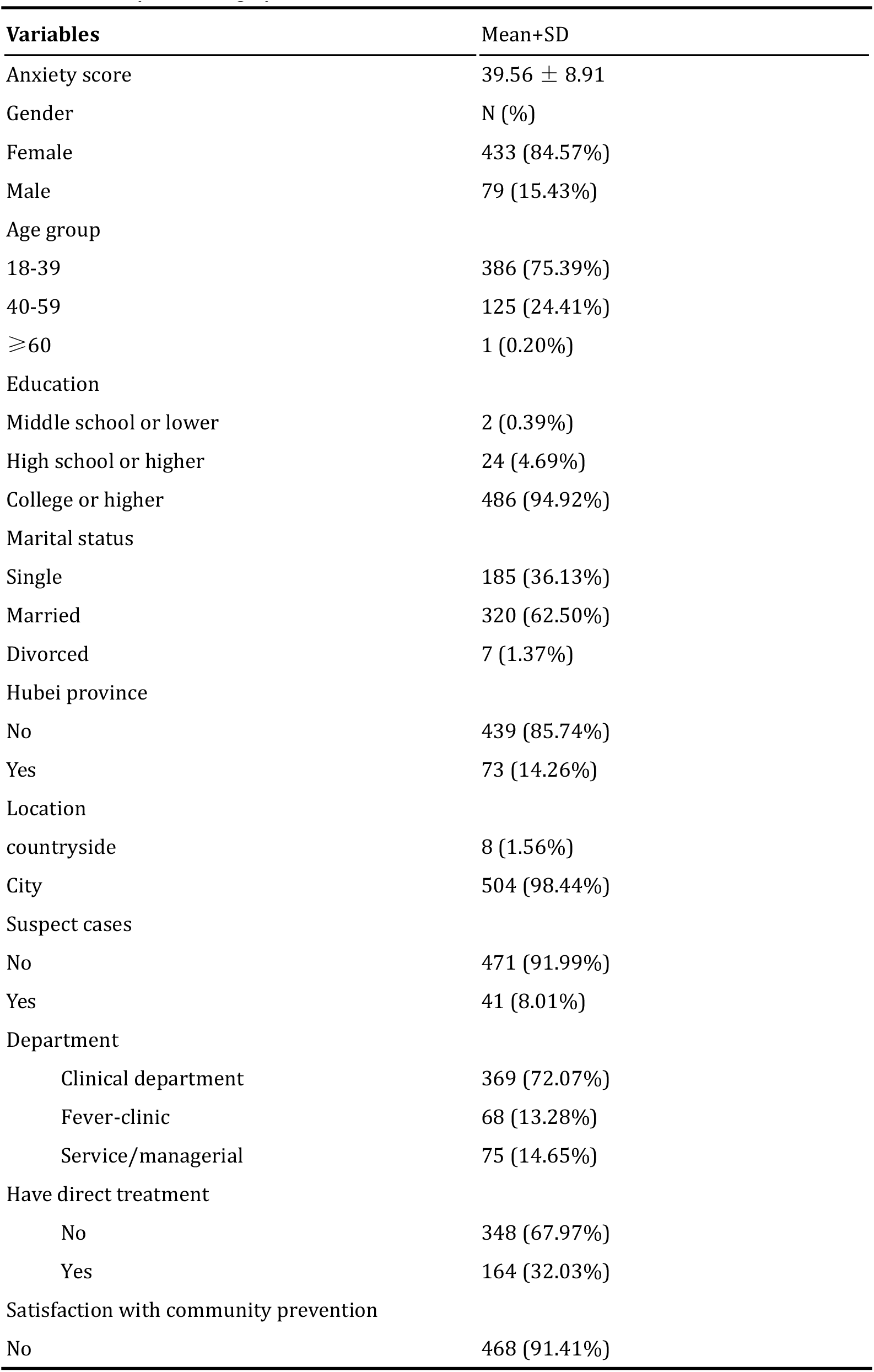
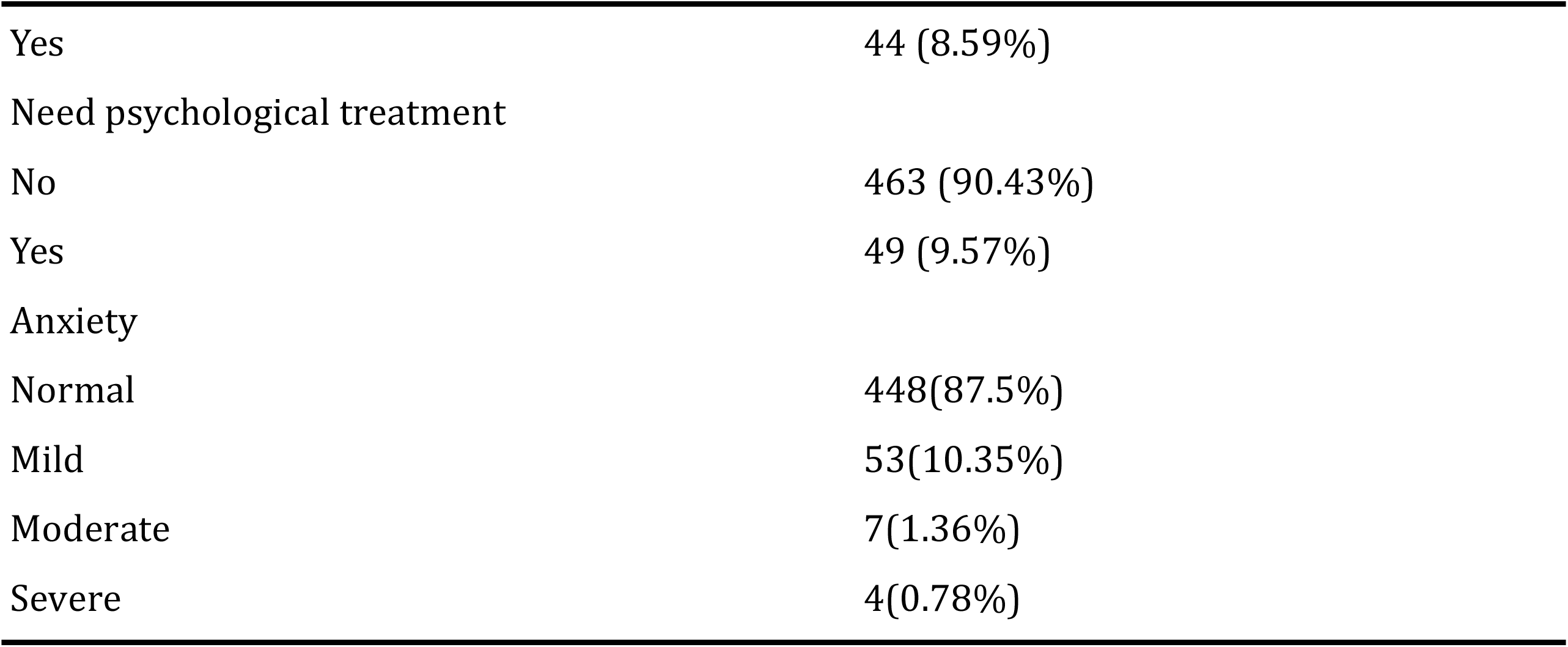
Participant demographic data

| **Variables** | **Mean±SD** |
| --- | --- |
| Anxiety score | 39.56 ± 8.91 |
| Gender | N (%) |
| Female | 433 (84.57%) |
| Male | 79 (15.43%) |
| Age group |  |
| 18-39 | 386 (75.39%) |
| 40-59 | 125 (24.41%) |
| ≥60 | 1 (0.20%) |
| Education |  |
| Middle school or lower | 2 (0.39%) |
| High school or higher | 24 (4.69%) |
| College or higher | 486 (94.92%) |
| Marital status |  |
| Single | 185 (36.13%) |
| Married | 320 (62.50%) |
| Divorced | 7 (1.37%) |
| Hubei province |  |
| No | 439 (85.74%) |
| Yes | 73 (14.26%) |
| Location |  |
| countryside | 8 (1.56%) |
| City | 504 (98.44%) |
| Suspect cases |  |
| No | 471 (91.99%) |
| Yes | 41 (8.01%) |
| Department |  |
| Clinical department | 369 (72.07%) |
| Fever-clinic | 68 (13.28%) |
| Service/managerial | 75 (14.65%) |
| Have direct treatment |  |
| No | 348 (67.97%) |
| Yes | 164 (32.03%) |
| Satisfaction with community prevention |  |
| No | 468 (91.41%) |
| Yes | 44 (8.59%) |
| Need psychological treatment |  |
| No | 463 (90.43%) |
| Yes | 49 (9.57%) |
| Anxiety |  |
| Normal | 448(87.5%) |
| Mild | 53(10.35%) |
| Moderate | 7(1.36%) |
| Severe | 4(0.78%) |

### Comparing the differences between two groups (direct treatment versus non-direct treatment)

**Table 2** shows the differences between medical staff who had directly treated patients with a confirmed case and those who had not. The average anxiety score was significantly higher in medical staff who directly treated confirmed cases, compared with those who did not (41.11 ± 9.79 versus 38.83 ± 8.38, p=0.007). There is a significant difference between these two groups in terms of the variables of gender, department, Hubei province, suspect case, satisfaction in terms of the effectiveness of community prevention, and the need for psychological counseling. However, the variables of age group, education, marital status and location were not obvious in these two groups.

**Table 2.**
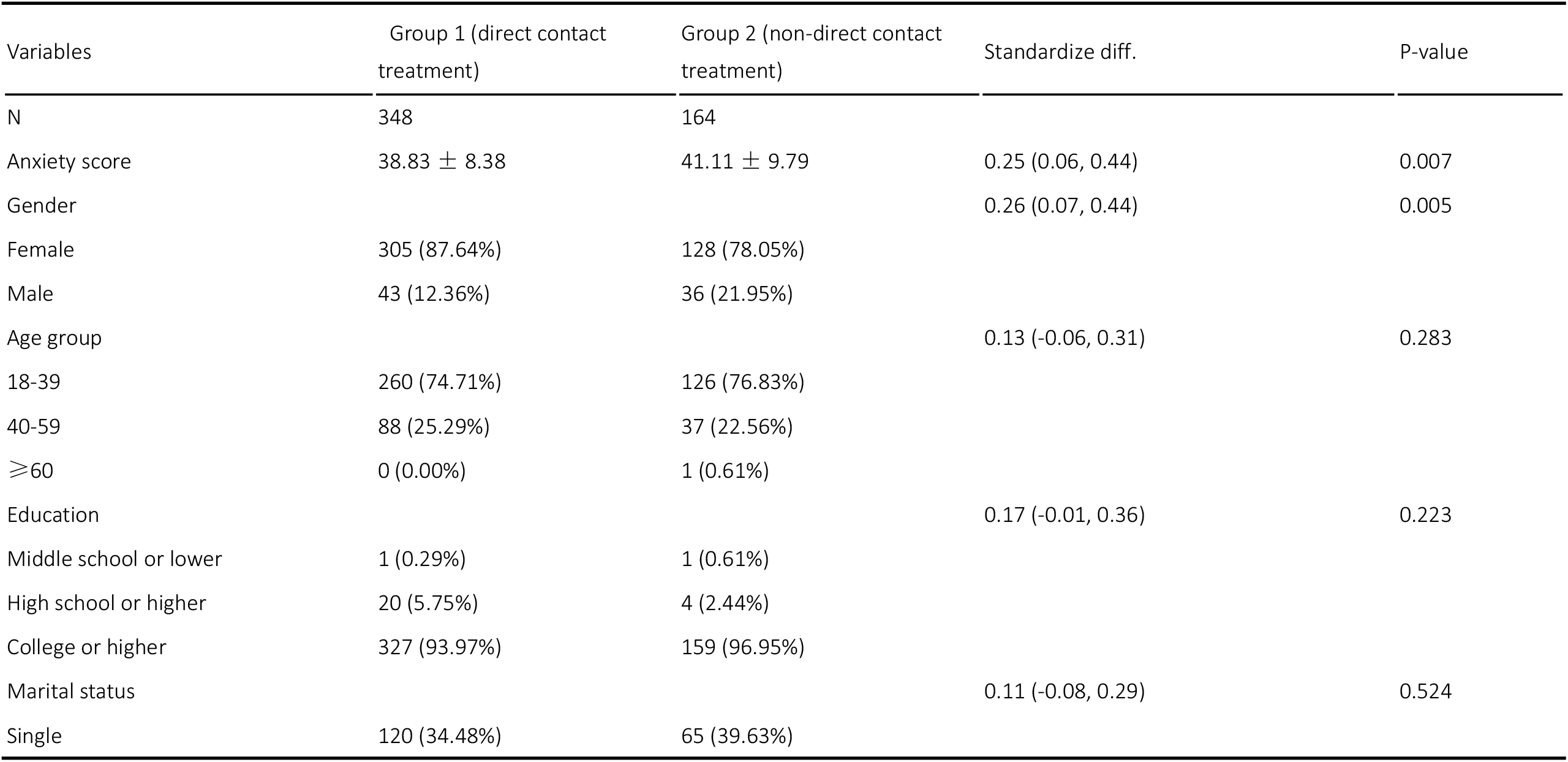
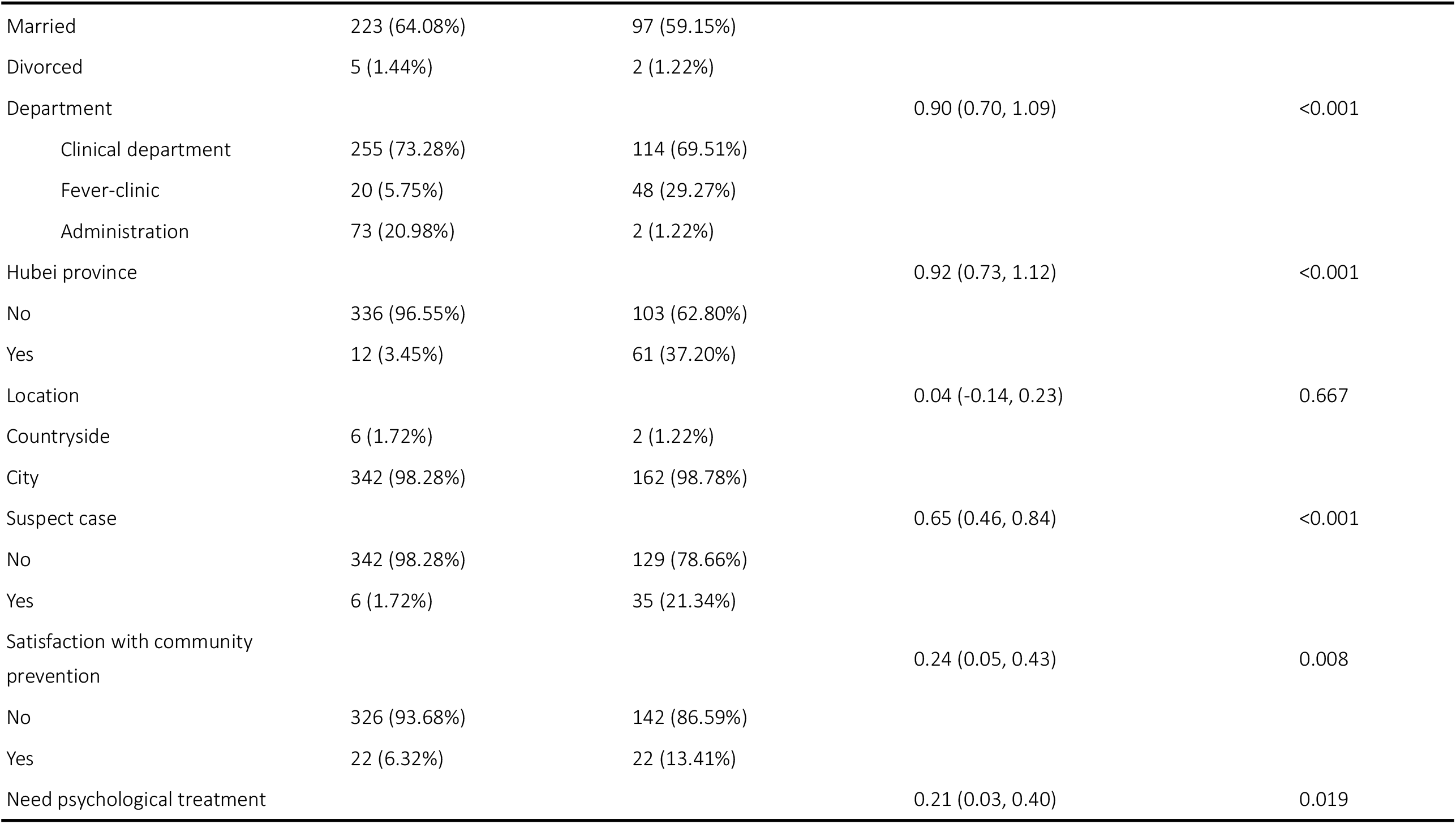
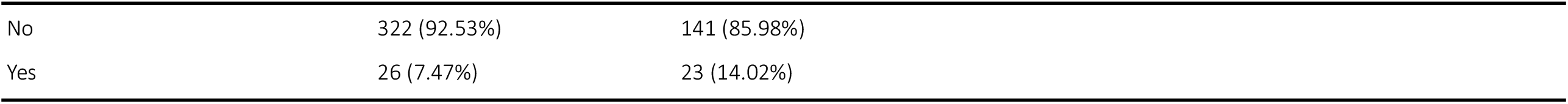
Comparing the differences between two groups (direct treatment versus non-direct treatment)

| Variables | Group 1 (direct contact treatment) | Group 2 (non-direct contact treatment) | Standardize diff. | P-value |
| --- | --- | --- | --- | --- |
| N | 348 | 164 |  |  |
| Anxiety score | 38.83 ± 8.38 | 41.11 ± 9.79 | 0.25 (0.06, 0.44) | 0.007 |
| Gender |  |  | 0.26 (0.07, 0.44) | 0.005 |
| Female | 305 (87.64%) | 128 (78.05%) |  |  |
| Male | 43 (12.36%) | 36 (21.95%) |  |  |
| Age group |  |  | 0.13 (-0.06, 0.31) | 0.283 |
| 18-39 | 260 (74.71%) | 126 (76.83%) |  |  |
| 40-59 | 88 (25.29%) | 37 (22.56%) |  |  |
| $\geq 60$ | 0 (0.00%) | 1 (0.61%) | | |
| Education |  |  | 0.17 (-0.01, 0.36) | 0.223 |
| Middle school or lower | 1 (0.29%) | 1 (0.61%) |  |  |
| High school or higher | 20 (5.75%) | 4 (2.44%) |  |  |
| College or higher | 327 (93.97%) | 159 (96.95%) |  |  |
| Marital status |  |  | 0.11 (-0.08, 0.29) | 0.524 |
| Single | 120 (34.48%) | 65 (39.63%) |  |  |
| Married | 223 (64.08%) | 97 (59.15%) |  |  |
| Divorced | 5 (1.44%) | 2 (1.22%) |  |  |
| Department |  |  | 0.90 (0.70, 1.09) | <0.001 |
| Clinical department | 255 (73.28%) | 114 (69.51%) |  |  |
| Fever-clinic | 20 (5.75%) | 48 (29.27%) |  |  |
| Administration | 73 (20.98%) | 2 (1.22%) |  |  |
| Hubei province |  |  | 0.92 (0.73, 1.12) | <0.001 |
| No | 336 (96.55%) | 103 (62.80%) |  |  |
| Yes | 12 (3.45%) | 61 (37.20%) |  |  |
| Location |  |  | 0.04 (-0.14, 0.23) | 0.667 |
| Countryside | 6 (1.72%) | 2 (1.22%) |  |  |
| City | 342 (98.28%) | 162 (98.78%) |  |  |
| Suspect case |  |  | 0.65 (0.46, 0.84) | <0.001 |
| No | 342 (98.28%) | 129 (78.66%) |  |  |
| Yes | 6 (1.72%) | 35 (21.34%) |  |  |
| Satisfaction with community prevention |  |  | 0.24 (0.05, 0.43) | 0.008 |
| No | 326 (93.68%) | 142 (86.59%) |  |  |
| Yes | 22 (6.32%) | 22 (13.41%) |  |  |
| Need psychological treatment |  |  | 0.21 (0.03, 0.40) | 0.019 |

|  |  |  |
| --- | --- | --- |
| No | 322 (92.53%) | 141 (85.98%) |
| Yes | 26 (7.47%) | 23 (14.02%) |

### Univariate analysis

#### Table 3 shows the risk factors associated with increased anxiety scores by using univariate analysis

The results reported that direct treatment, being from Hubei province and suspect cases were associated with increased anxiety scores (*P* ≤ 0.05). However, we found that the variables in terms of gender, age, education, marital status, location, healthcare workers’ satisfaction with the effectiveness of community prevention measures, and the need for psychological counseling, did not increase anxiety scores (all *P* ≥ 0.05)

**Table 3.**
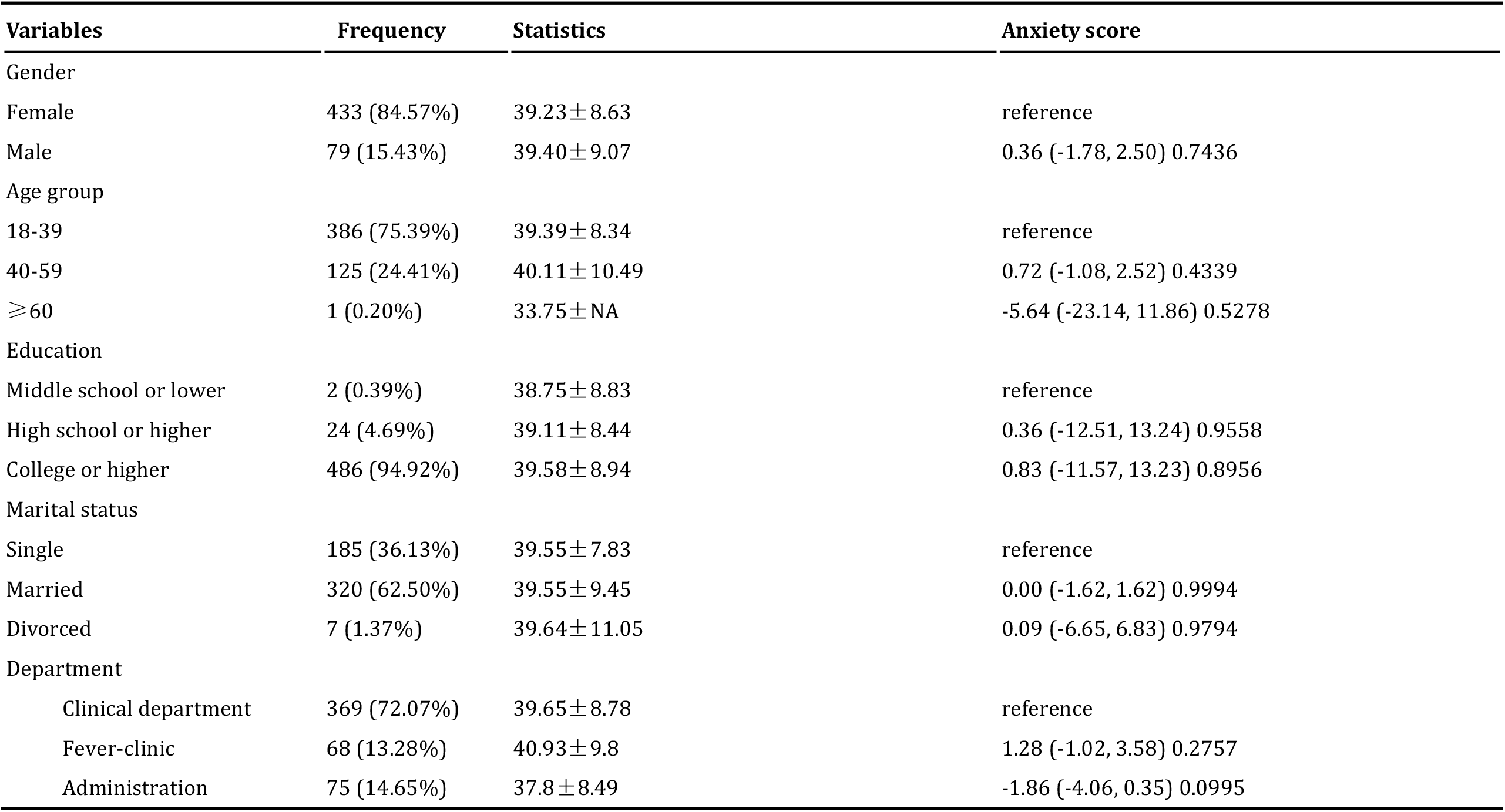
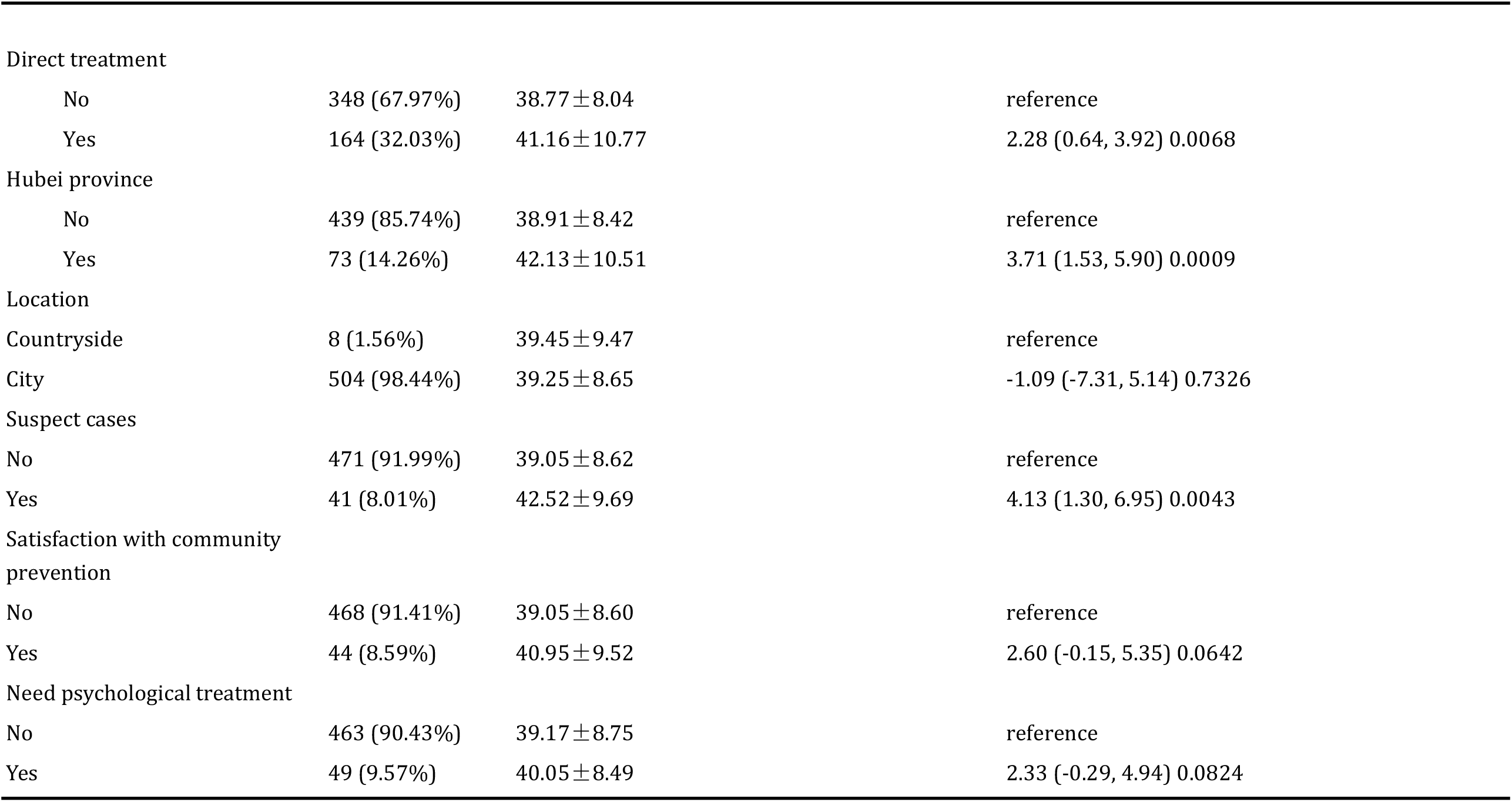
shows the risk factors associated with increased anxiety scores by using univariate analysis

### Multivariable analysis to evaluate the independent impact of direct treatment on anxiety scores among medical workers using non-adjusted and adjusted linear regression analysis

We used multivariable linear regression to detect the relationship between direct treatment and anxiety score using a different model. The results show that direct treatment is an independent risk factor for an increased anxiety score (*β*value =2.280 95%CI: 0.636-3.924; p=0.0068) in an unadjusted model, together with suspect cases (*β*value =4.13 95%CI: 1.30-6.95; p=0.0043), and being a medical worker from Hubei province (*β*value =3.71 95%CI: 1.53-5.90; p=0.0009). In addition, all of these associations were still significant after adjusting for the variables of gender, age, education and marital status, with detailed results shown in Table 4.

**Table 4.** Multiple linear regression for anxiety score

| Variables(exposure) | Non-adjusted( $\beta$ ,95%,CI,*P*) | Adjusted( $\beta$ ,95%,CI,*P*) |
| --- | --- | --- |
| Direct contact treatment |  |  |
| No | reference | reference |
| Yes | 2.28 (0.64, 3.92) 0.0068 | 2.33 (0.65, 4.00) 0.0068 |
| Suspect cases |  |  |
| No | reference | reference |
| Yes | 4.13 (1.30, 6.95) 0.0043 | 4.44 (1.55, 7.33) 0.0028 |
| Hubei province |  |  |
| No | reference | reference |
| Yes | 3.71 (1.53, 5.90) 0.0009 | 3.67 (1.44, 5.89) 0.0013 |

Outcome: anxiety score
Non-adjusted model adjusted for: We did not adjust for other covariants
Adjusted model adjusted for: Gender, age, education, marital status
Ref: reference

## Discussion

Our study examined the prevalence of anxiety in medical staff and identified their risk factors for increased anxiety. Our results found that working in COVID-19 patient care, suspect cases and being from Hubei province were all risk factors for increased anxiety scores, which is an important finding suggesting that government authorities need to implement measures to alleviate mental health symptoms early on.

New bio-disasters - such as SARS, Ebola, H1N1, Middle East Respiratory Syndrome and the Coronavirus - are profoundly associated with adverse psychological effects on medical staff, including depression, anxiety and insomnia. Using SAS, 12.5% of medical staff experienced anxiety, which is less than in a previous study that reported stress reactions during the SARS outbreak^16^. The main possible reasons were the different tool assessments. By the time of our research study, almost four weeks after the first COVID-19 patient was confirmed in Wuhan, China at the end of December 2019, health workers apparently had sufficient time to adjust themselves to caring for infected patients and feeling confident in doing so. In addition, government and healthcare authorities provided stricter and safer protective measures to support them, which to some extent reduced workers’ anxiety, in contrast to the early peak of the epidemic. However, Hyunsuk Jeong’s study reported the prevalence of anxiety symptoms in people who were not diagnosed with MERS and needed two weeks of isolation was 7.6% (95% confidence interval [CI], 6.3% to 8.9%), which is less than in our study^17^. The discrepancy might be due to the fact there were only 267 health workers (16.1%) and the authors used the 7-item Generalized Anxiety Disorder Scale to assess anxiety, with a cut-off of 5 points confirming mild anxiety. Nevertheless, medical workers who provided direct treatment or care for infected patients suffered higher anxiety scores, compared to those who did not care for COVID-19 patients. Previous studies have reported that psychological symptoms, such as anxiety, depend on the epidemic phase^18^. This is because medical workers might experience psychological adaptation after gradually learning more about SARS and obtaining rich clinical experience in the treatment and care of infected patients.

The results show that health workers from Hubei, the most severely affected area, had higher anxiety scores (β value =3.71, CI: 1.53-5.90; *P*= 0.0009) compared to health workers from other places. Staff working in hospitals in Hubei have suffered heavy workloads due to the increasing number of infected cases that require centralization to designated hospitals for standard isolation treatment. Additionally, the media have reported that medically protective materials, such as N95 masks, goggles and protective clothing were severely deficient during the early stages of the outbreak^19^. All of these factors invisibly aggravated the psychological burden.

In addition, our results found that staff with suspect infection cases had higher anxiety scores than non-suspect cases (**β**value=4.44, CI: 1.55 −7.33; *p=*0.0028). Ping Wu and colleagues^9^ conducted a survey on the psychological problems of hospital employees during the SARS epidemic, and found that respondents who were quarantined had higher levels of posttraumatic stress than those without exposure. Medical staff became suspect cases mainly through hospital-related transmission. Suspect cases with a high risk of infection needed to remain isolated for two weeks under medical observation^20^. During that time, they might suffer a dilemma: on the one hand, most people fear becoming infected with SARS-CoV-2, while on the other hand, they also struggle with not taking on responsibility for fighting COVID-19. They also worry about the health risks to their own family, leading to a psychological burden. This complex situation might further intensify anxiety in medical staff. Therefore, governments should focus on potential psychological problems among suspect cases in medical staff, and provide effective mental health measures to alleviate suffering.

After adjusting the confounding factors, providing direct treatment to infected patients was an independent factor in increased anxiety scores, compared to not directly caring for COVID-19 patients. Participating in treatments or procedures for infected patients was a challenging job for frontline staff who experienced stress, as they were at high potential risk of infection due to the illness’ characteristics of high transmission efficiency, rapid deterioration and pathogenicity. Hence, healthcare workers, such as those having direct contact with patients, suffered higher anxiety symptom scores than staff who were at low risk. Therefore, although the Chinese government and society compliment medical personnel for their dedication in fighting COVID-19, which could encourage medical workers and make them feel honored and proud to participate in this difficult mission, authorities should also focus on implementing measures to target workers’ mental health. As the SARS-Cov-2 epidemic is becoming a global issue, fighting COVID-19 appears to be a sustained event, which might result in medical personnel suffering psychological problems.

Our study has some limitations. First, the questionnaires were dispatched non-randomly via WeChat, so a selective-bias exists in our study, and the number of medical staff from Hubei was in a minority (14.26%), which means that our study does not completely reflect the entire mental health picture of Chinese medical staff in quarantine. Second, we did not collect data during the early stages of the epidemic, when anxiety score levels could have been different. A previous study reported that the prevalence of psychological disorders presented differently from the start to the end of the outbreak^18^. Third, our study used a cross-sectional design that cannot determine causality for factors and outcome. In addition, a comprehensive assessment, including demographic factors, such as years of experience, having children or not and a history of mental disorders, would be beneficial in analyzing potential anxiety factors. However, our study also has some strengths. To our knowledge, this is the first study to assess anxiety levels among medical staff in China during the period of COVID-19, and we used comprehensive data statistical analyses to make our results reliable.

## Conclusion

Our study found the prevalence of anxiety is mild, however, medical staff who had had direct contact through treatment of infected patients may experience an increase in their anxiety score, compared to workers who had not had direct contact with infected patients. In addition, healthcare workers who were quarantined in Hubei province, and those who were suspect cases also saw increased anxiety scores. Therefore, government and healthcare authorities should proactively implement appropriate measures, such as providing psychological counselling services, to prevent, alleviate or treat increased anxiety among medical staff during the COVID-19 epidemic.

## Data Availability

The data can be obtained by some research applying for study

https://mail.163.com/js6/main.jsp?sid=SCLpuBPuhWzbsqzilduuykMyEaqvfhKe&df=mail163_letter#module=read.ReadModule%7C%7B%22area%22%3A%22normal%22%2C%22isThread%22%3Afalse%2C%22viewType%22%3A%22%22%2C%22id%22%3A%2288%3A1tbiWBreY1uHrLFbvAAAsZ%22%2C%22fid%22%3A1%7D

## Author Contributions

Xiao-Ming Zhang and Chenyun Liu had full access to all study data and take responsibility for data integrity and accuracy of the data analysis.

Concept and design: Xiao-Ming Zhang

Data acquisition, analysis or interpretation: XinYing-Xu and Yunzhi-Yang Drafting of the manuscript: Chenyun-Liu and Xiao-Ming Zhang

Administrative, technical or material support: Qing-Li Dou and Wen-Wu Zhang

## Acknowledgements

The authors thank all frontline medical staff who are fighting COVID-19. We sincerely appreciate their participation in our study.

